# Assessing the feasibility and acceptability of a bespoke large language model pipeline to extract data from different study designs for public health evidence reviews

**DOI:** 10.1101/2025.07.21.25331917

**Authors:** Zalaya Simmons, Beti Evans, Tamsyn Harris, Harry Woolnough, Lauren Dunn, Jonathon Fuller, Kerry Cella, Daphne Duval

## Abstract

**Introduction:** Data extraction is a critical but resource-intensive step of the evidence review process. Whilst there is evidence that artificial intelligence (AI) and large language models (LLMs) can improve the efficiency of data extraction from randomised controlled trials, their potential for other study designs is unclear. In this context, this study aimed to evaluate the performance of a bespoke LLM model pipeline (Retrieval-Augmented Generation pipeline utilising LLaMa 3-70B) to automate data extraction from a range of study designs by assessing the accuracy, reliability and acceptability of the extractions.

**Methods:** Accuracy was assessed by comparing the LLM outputs for 173 data fields with data extracted from a sample of 24 articles (including experimental, observational, qualitative, and modelling studies) from a previously conducted review, of which 3 were used for prompt engineering. Reliability (consistency) was assessed by calculating the mean maximum agreement rate (the highest proportion of identical returns from 10 consecutive extractions) for 116 data fields from 16 of the 24 studies. Acceptability of the accuracy and reliability outputs for each data field was assessed on whether it would be usable in real-world settings if the model acted as one reviewer and a human as a second reviewer.

**Results:** Of the 173 data fields evaluated for accuracy, 68% were rated by human reviewers as acceptable (consistent with what is deemed to be acceptable data extraction from a human reviewer). However, acceptability ratings varied depending on the data field extracted (33% to 100%), with at least 90% acceptability for ‘objective’, ‘setting’, and ‘study design’, but 54% or less for data fields such as ‘outcome’ and ‘time period’. For reliability, the mean maximum agreement rate was 0.71 (SD: 0.28), with variation across different data fields.

**Conclusion:** This evaluation demonstrates the potential for LLMs, when paired with human quality assurance, to support data extraction in evidence reviews that include a range of study designs, however further improvements in performance and validation are required before the model can be introduced into review workflows.

## 1. Introduction

The advancement of artificial intelligence (AI) offers new opportunities to improve the efficiency and scalability of evidence reviews by accelerating and automating steps in the review process (1). Traditional machine learning (ML) approaches, which involve training algorithms on structured and task-specific datasets, may already be integrated into the evidence review process through several methods, such as the use of machine learning algorithms to prioritise relevant articles in the title and abstract screening process (2, 3).

The emergence of generative large language models (LLMs), such as GPT, Claude and LLaMa, has introduced further opportunities for automation of steps in the evidence review process. LLMs are pre-trained on large quantities of unstructured text which enables them to follow instructions in natural language and be applied more flexibly across a wider range of tasks (4). This can include extracting textual and numerical data from full text articles (5) or drafting text for reports (6).

Generative LLMs have shown promising performance for some steps of the evidence review process, such as screening (7) and data extraction (8–10). However, results have been less promising for other steps such as search strategy generation (11), or risk of bias assessments (12). Additional limitations such as hallucinating content, misinterpreting or oversimplifying nuanced data, and generating outputs which can be difficult to validate, raise concerns around transparency, reproducibility and the integrity of outputs produced by generative LLMs (1).

Of the steps in the evidence review process that have potential for AI integration, data extraction can be particularly resource and time-intensive, and requires a high degree of accuracy to uphold validity (13). There is the potential to use LLMs alongside human reviewers to semi-automate this task to improve efficiency. However, while LLMs have shown promising results when extracting data from randomised controlled trials (RCTs) (8–10, 14), their ability to extract data from a wider range of study designs, including observational research, is unclear. This is particularly important in public health, where evidence is often synthesised from diverse study designs to inform advice, guidance, policy and practice. To address this gap, data scientists and evidence reviewers at

UKHSA co-developed a bespoke LLM pipeline for semi-automated data extraction for evidence reviews (evidence synthesis outputs following systematic methodologies), integrated within UKHSA infrastructure and designed to work with human oversight.

The aim of this feasibility study was to evaluate the performance of a bespoke LLM pipeline designed to automate data extraction in evidence reviews and assess the accuracy, reliability and acceptability of outputs to human reviewers.

## 2. Methods

A Retrieval-Augmented Generation (RAG) pipeline utilising a LLM was developed to automate data extraction (see Figure 1). The LLM used was LLaMa 3-70B.

**Figure 1.**
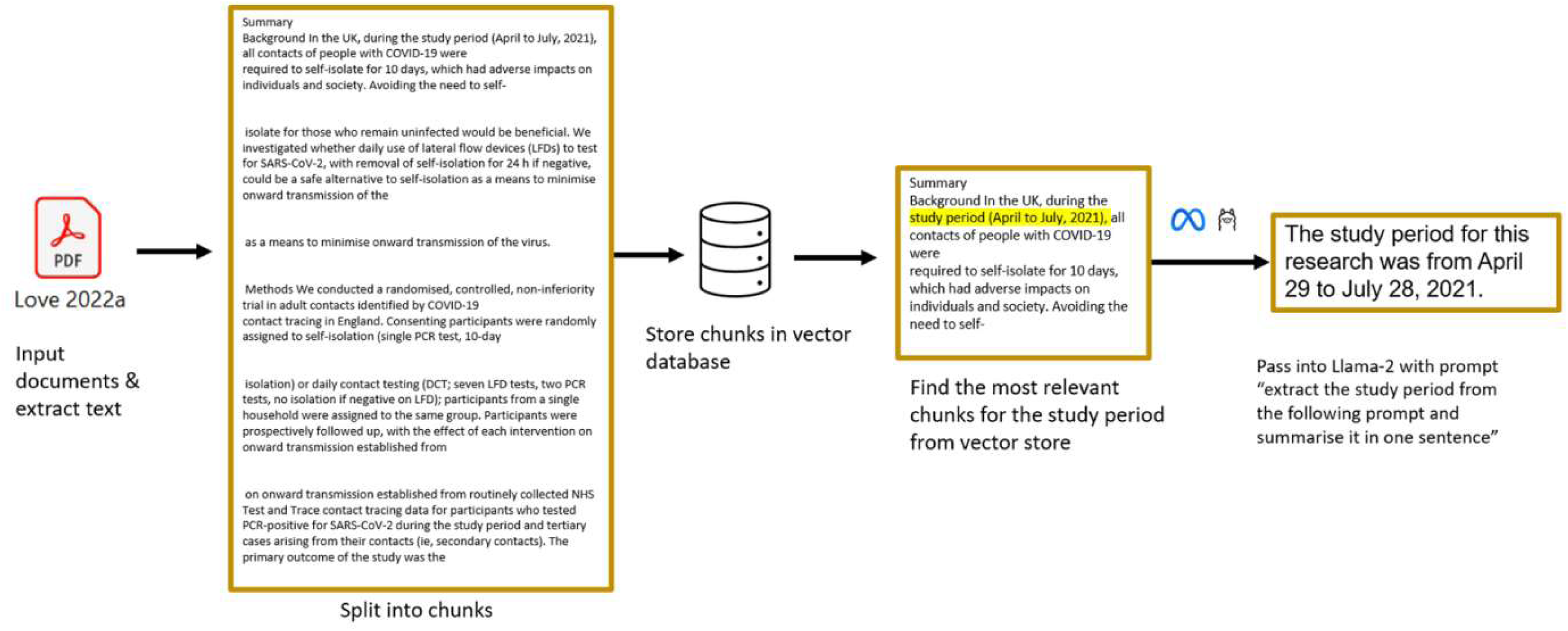
RAG Pipeline

### 2.1 Reference standard

LLM data extraction outputs were compared against data that had been extracted and quality assured by 2 reviewers as part of a previously published rapid mapping review (15, 16). The rapid mapping review aimed to identify and categorise the evidence generated during the COVID-19 pandemic on the effectiveness of non-pharmaceutical interventions (NPIs) implemented in community settings in the UK.

A convenience sample of 24 open access studies was selected from the 151 studies included in the review, ensuring the sample contained examples of the different study designs included (2 RCTs, 5 prospective longitudinal, 3 cross-sectional, 4 ecological, 4 mixed-methods, 3 qualitative, and 3 modelling studies) (17–40). This was to ensure that the model’s performance in extracting data from different study designs that are regularly encountered when reviewing evidence relevant to public health could be evaluated. PDF copies of the articles were accessed within the organisation’s secure infrastructure, with no data used to train the LLM, no third party access permitted, and no data leaving the internal environment.

### 2.2 Data fields extracted

The LLM extracted a total of 173 data fields from 24 full text articles, which corresponded to those that had been extracted by human reviewers for the published rapid mapping review (15, 16). The data fields extracted varied by study design. Seven data fields were extracted for each study design except for RCTs and modelling studies for which 8 data fields were extracted (see Table 1 for details of data fields extracted).

**Table 1.** Data fields extracted and prompts used.

| Data field | Study designs for which data field extracted | Data field prompt used in the model | Expected extraction | Example human reviewer extraction |
| --- | --- | --- | --- | --- |
| Study objective | All study designs (RCTs, prospective longitudinal, cross-sectional, ecological, mixed-methods, qualitative, modelling) | ‘What was the objective of the study?’ | Summary of the study’s objective | To assess whether daily testing of COVID-19 contacts in secondary schools resulted in a similar control of COVID-19 transmission and increased school attendance compared to self-isolating school-based COVID-19 contacts |
| Study design | All study designs except modelling studies | ‘What was the study design?’ | Named study design | Open-label, cluster randomised controlled trial |
| Setting | All study designs | ‘What was the setting that the study was set in? (country, city, or specific location)’ | Setting such as schools, community-wide, and or location/nation if applicable | England (secondary schools and further education colleges) |
| Participants/ population* | All study designs except modelling studies | ‘What were the number of participants and groups used in the study?’ | Participant numbers and relevant demographics<br>For ecological studies, details of the population included in the study | Control group: n=99 schools (n=102,859 students, n=11,798 staff); students: 47.1% aged 11 to 14 years, 48.1% aged 15 to 18 years<br>Intervention group: n=102 schools (n=111,693 students, n=12,229 staff); students: 45.1% aged 11 to 14 years, 46.7% aged 15 to 18 years |
|  |  |  |  | No restriction on age: students aged 19 years or more attended further education colleges (less than 0.1% in both the control and intervention group) |
| Study period | All study designs | 'What was the duration of the study? (e.g. 10 weeks)' | Time period (dates) | 10-week study starting between 19 April to 10 May 2021, to 27 June 2021 |
| NPI | All study designs | 'What are the non-pharmaceutical interventions this study assessed and how were they used?' | Named NPI(s) evaluated in the study | Test and release strategies |
| Outcomes | All study designs | 'What were the outcomes this study monitored?' | Outcomes evaluated in the study relevant to review question | COVID-19 transmission (rate of symptomatic polymerase chain reaction [PCR]-positive infection, controlled for community case rates)<br>Lost time (school or work) |
| Control group | RCTs | 'What was the control group used in the study?' | Details of the control used in experimental studies | Students or staff contacts of lateral flow device or PCR positive COVID-19 cases self-isolated for 10 days |
| Model | Modelling studies | 'What was the model used in this study?' | Type of model used | An agent-based model |
| Data | Modelling studies | 'What was the data used in the modelling study?' | Details of data sources used to populate the model | Synthetic population of 103,000 generated from 2011 UK Census data |
| Scenarios | Modelling studies | 'What were the simulated scenarios used in this study?' | Outline of scenarios used in the model | Simulate the impact on viral spread of various combinations of:<br>• proportion of contact tracing application (CTA) users in the population<br>• levels of testing capacity<br>• levels of compliance with self-isolation on the part of CTA users<br>• testing policy |

\*'population' for ecological studies and 'participants' for other study designs

### 2.3 Pipeline development and prompt engineering

A RAG pipeline utilising an LLM (LLaMa 3-70B) was developed to automate data extraction (Figure 1). PDF copies of the articles for data extraction were inputted into the pipeline. Text from each PDF was tokenized (broken down into smaller units [tokens]), split into chunks and stored in a vector database (a type of database that stores text as numbers based on meaning, so similar pieces of text can be found quickly) using Python and the Langchain framework. The chunking of data is necessary to enable processing within the context window, which equates to the maximum amount of data that can be passed to the LLM at one time. These chunks of text are then turned into numbers (called vectors), that represent their meaning so that the model can understand and compare them. This permits the database to be queried and select the most relevant chunks to the user’s query. The chunks were then selected for each data field, from which the LLM extracted and summarised the information using prompting. The summarised information was outputted into an Excel file (see example in supplementary material Table S1).

Prompts for each data field extracted were developed in an iterative and collaborative process between evidence reviewers and data scientists using 4 studies (17, 36, 39, 41), 3 of which were included in the evaluation (17, 36, 39). For the purpose of generating consistent responses with a rich vocabulary, the temperature setting of the model (which controls randomness) was set very low (0.01) to reduce randomness and ensure consistency, and the top_p setting (which controls how much of the vocabulary was considered) was set high to (0.99). Table 1 shows the prompts used in the LLM and examples of expected extracted content for each data field. See supplementary material Table S2 for system message and prompt for the overall task.

### 2.4 Evaluation

An evaluation framework was developed to assess the accuracy, reliability, and acceptability of the LLM’s outputs for each data field based on error classification used in a published evaluation (8). Using these frameworks, 2 reviewers (TH, BE) independently assessed the outputs for each data field according to the pre-defined error and acceptability criteria (Tables 2 and 3). Assessments were then agreed by consensus, with a third reviewer (ZS) present to resolve disagreements and record decisions.

**Table 2.** Accuracy evaluation framework.

| Error | Description | Acceptability criteria |
| --- | --- | --- |
| No error | The model returns the correct answer | Extraction **acceptable** |
| Major error | The data is completely misrepresented in a way that significantly impairs the integrity of the review, for example including interventions or outcomes that were not evaluated in the article | All major errors are **unacceptable** |
| Minor error | Involves slight misrepresentation of data that, while inaccurate, does not impact the review's results. This also includes cases where small but non-critical data is missing. For example, for a study conducted in Glasgow, Scotland, the model response is that the study was conducted in a city in Scotland | Minor errors are **acceptable** if they would be an acceptable mistake a human would make, for example misspelling a word or missing off a small part of a phrase.<br>**Unacceptable** errors are where a human would be unlikely to make that mistake, or data is misrepresented enough there is potential for it to be misinterpreted |
| Missing data | The model has not extracted data (whether value not returned, or declared that there was not relevant data in the chunks) | All missing data is **unacceptable** if there is data available in the article and a human reviewer would have been able to extract |
| Not stated in article | The information of interest has not been stated in the source article (for example, a study design classified as ecological by a human reviewer but 'ecological' is not stated in the source article) | **Acceptable** if the model has been able to infer the correct response, or correctly state that the data was not available in the article.<br>**Unacceptable** if the model has presented incorrect data from another part of the article |
| Hallucination | The model has invented data that is not from anywhere in the source article | Hallucination is always **unacceptable** |

**Table 3.** Reliability evaluation framework.

| Error | Description | Acceptability criteria |
| --- | --- | --- |
| No error | The model presents the same information for all returns | Errors are **unacceptable** if variations in data extractions lead to fundamentally different understandings or interpretations of the data. All 'value not returned' errors are **unacceptable**.<br>Errors are **acceptable** if, across 10 extractions, the information conveyed is essentially the same, allowing for consistent interpretation despite minor variations in wording or formatting. |
| Value error | The model presents different information (values) across the 10 returns. For example:<br>• There were 10 male participants (1 return)<br>• There were 11 male participants (1 return)<br>• There were 10 participants (8 returns) |  |
| Voice error | The model presents the same information (values) in different formats across multiple returns (for example, for every 10 returns, 6 are returned as paragraphs and 4 are returned as bullet points) or has minor variation in wording (for example change in tense such as 'The number of participants and groups used in the study were...' or 'The number of participants and groups used in the study are...') |  |
| Value not returned | The model has not extracted data |  |

### Accuracy

Accuracy of the LLM’s output for each data extraction field for the 24 studies was assessed according to the evaluation framework (Table 2). Each data field was assigned one error type (no error, major, minor, missing data, not stated in article, or hallucination). The LLM outputs were compared to the data extracted by human reviewers for the published mapping review, defining accuracy as the proportion of extractions with no errors.

### Reliability

Reliability (consistency) of outputs produced by the LLM was assessed by running 10 extractions consecutively using identical prompts. The reference standard for reliability was that the LLM was expected to return identical values in the same format. The group of 10 extractions for each data field was evaluated against pre-defined error categories (no error, value error, voice error, or value not returned) (Table 3). Each data field could be designated as having both a value and voice error, or one error category only. Reliability was assessed for each data field for 16 studies (2 RCTs, 3 prospective longitudinal, 1 cross-sectional, 4 ecological, 2 mixed-methods, 2 qualitative, 2 modelling) (17–19, 21–28, 31, 33, 34, 36, 39).

### Acceptability

Acceptability of both accuracy and reliability outputs produced by the LLM was assessed against whether the output was useable, and whether the level of variation would be accepted in a human reviewer’s work (Tables 2 and 3). Errors could be acceptable if they were consistent with the types of errors a human reviewer might reasonably make (such as minor differences in wording), and did not affect interpretation of the study findings. Errors were considered unacceptable if they risked misinterpretation of the study characteristics or findings.

### Analysis

Results were analysed in Python 3.11 using the Pandas library for data wrangling (also called data cleaning or preprocessing) and the Matplotlib and Plotly libraries for visualisation. Distribution of error types and the acceptability of identified errors were analysed for extractions both overall and disaggregated by data field and study design. Maximum agreement rate (the highest proportion of the 10 extractions that returned identical extractions) was used as a measure of reliability.

## 3. Results

### 3.1 Accuracy

The accuracy of 173 data fields extracted from 24 full text articles was assessed. Findings are presented in Table 4 and Figure 2. Of the 173 data fields extracted, 42% of outputs were accurate (no error), 28% had minor errors, 21% had major errors, 4.6% were categorised as missing data, 3.5% as data not stated in article, and 1.2% of errors were categorised as hallucinations. Overall, 68% of outputs returned were designated acceptable (consistent with what is deemed to be acceptable data extraction from a human reviewer).

**Figure 2.**
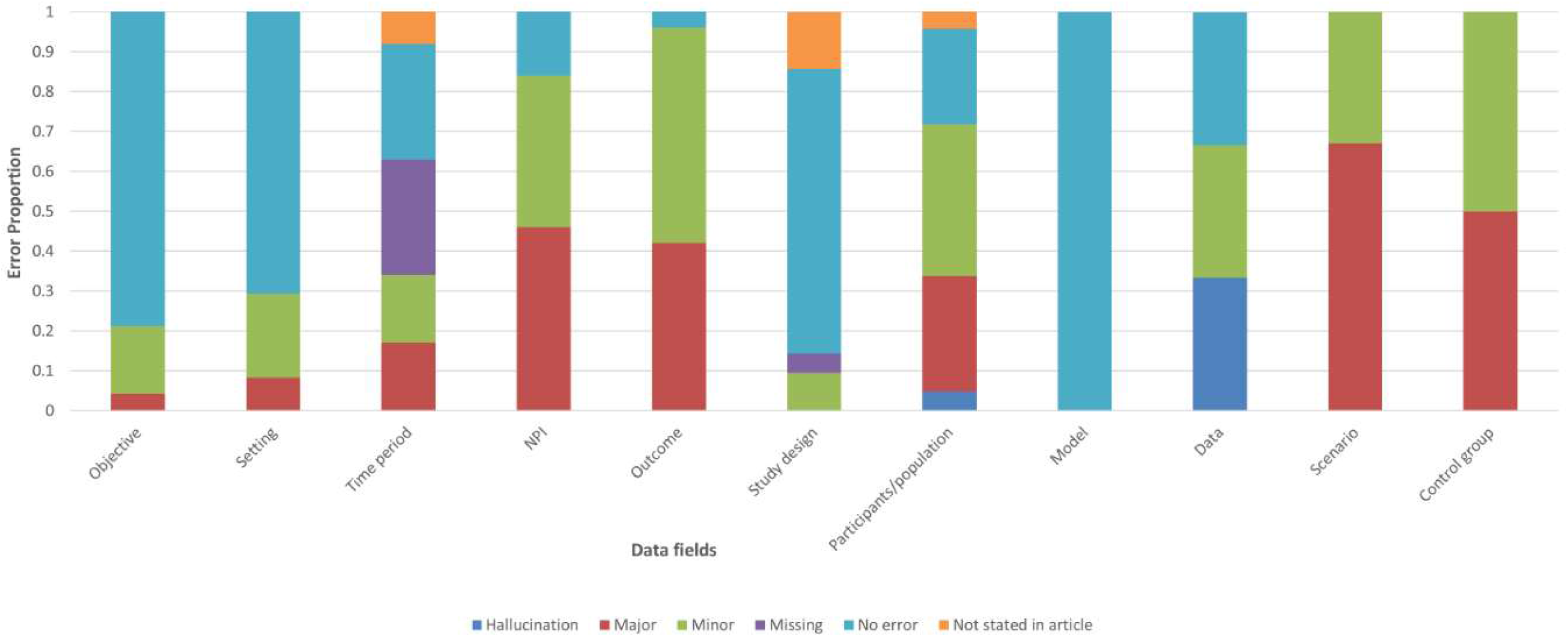
Distribution of accuracy error types by data field

**Table 4.** Accuracy errors and acceptability by data field (total number of extractions = 173)

| **Data field**<br>**(number of**<br>**studies)** | **No error**<br>**(%)** | **Minor**<br>**(%)** | **Major**<br>**(%)** | **Missing**<br>**(%)** | **Not stated in**<br>**article (%)** | **Hallucination**<br>**(%)** | **Acceptability**<br>**(% acceptable)** |
| --- | --- | --- | --- | --- | --- | --- | --- |
| Objective (24) | 19 (79%) | 4 (17%) | 1 (4.2%) | 0 | 0 | 0 | 22 (92%) |
| Setting (24) | 17 (71%) | 5 (21%) | 2 (8.3%) | 0 | 0 | 0 | 22 (92%) |
| Time period (24) | 7 (29%) | 4 (17%) | 4 (17%) | 7 (29%) | 2 (8.3%) | 0 | 13 (54%) |
| NPI (24) | 4 (17%) | 9 (38%) | 11 (46%) | 0 | 0 | 0 | 11 (46%) |
| Outcome (24) | 1 (4.2%) | 13 (54%) | 10 (42%) | 0 | 0 | 0 | 12 (50%) |
| Study design (21) | 15 (71%) | 2 (9.5%) | 0 | 1 (4.8%) | 3 (14%) | 0 | 19 (90%) |
| Participants/<br>population (21) | 5 (24%) | 8 (38%) | 6 (29%) | 0 | 1 (4.8%) | 1 (4.8%) | 14 (67%) |
| Model (3) | 3 (100%) | 0 | 0 | 0 | 0 | 0 | 3 (100%) |
| Data (3) | 1 (33%) | 1 (33%) | 0 | 0 | 0 | 1 (33%) | 1 (33%) |
| Scenario (3) | 0 | 1 (33%) | 2 (67%) | 0 | 0 | 0 | 0 |
| Control group (2) | 0 | 1 (50%) | 1 (50%) | 0 | 0 | 0 | 1 (50%) |
| **Totals** | **72 (42%)** | **48 (28%)** | **37 (21%)** | **8 (4.6%)** | **6 (3.5%)** | **2 (1.2%)** | **118 (68%)** |

However, findings varied across data fields, with acceptability ranging from 33% to 100% (Table 4). For data fields where at least 21 studies were evaluated, the proportion of data fields with no error ranged from 4.2% for ‘outcome’ to 79% for ‘objective’, whilst the proportion with a major error ranged from 0 for ‘study design’ to 46% for ‘NPI’. At least 90% of the outputs for data fields ‘objective’, ‘setting’, and ‘study design’ were designated acceptable by human reviewers (with a high no error rate of more than 70%). The data fields ‘NPI’ and ‘outcome’ were designated acceptable for 50% or less of extractions. The data field ‘model’ (for modelling studies only) had a correct extraction (no error) rate and acceptability of 100%, however this was only assessed in 3 studies.

Accuracy acceptability findings also varied by study design, from 56% overall for RCTs (2 studies) to 86% for cross-sectional studies (3 studies) (supplementary material Table S3). Interestingly, the acceptability of the outputs for the data field’s ‘objective’ and ‘setting’ was 100% for all study designs, except for mixed methods studies (4 studies, 75% for both data fields) and for qualitative studies (3 studies, 67% for both data fields). However, the number of studies in each study design category was too low to allow conclusions about differential model performance by study design to be drawn.

### 3.2 Reliability

The reliability of 116 data fields extracted from 16 full text articles was assessed. The model’s reliability (consistency) varied across different data fields. The mean maximum agreement rate across all data fields was 0.71 (SD: 0.28), showing that on average 7.1 of the 10 consecutive extractions returned the same value for a given data field (Table 5). The data fields ‘study design’, ‘setting’, and ‘objective’ showed higher consistency, with mean maximum agreement rates above 0.85. Lower agreement rates were observed for fields such as ‘outcome’ (mean: 0.55), and ‘participants/population’ (mean: 0.61).

**Table 5.** Reliability evaluation maximum agreement rate.

| **Data field**<br>**(number of studies)** | **Maximum agreement rate**<br>**(mean, SD)** |
| --- | --- |
| Objective (16) | 0.86 (0.23) |
| Setting (16) | 0.86 (0.19) |
| Time period (16) | 0.69 (0.26) |
| NPI (16) | 0.52 (0.25) |
| Outcome (16) | 0.55 (0.26) |
| Study design (14) | 0.94 (0.15) |
| Participants/population (14) | 0.61 (0.22) |
| Model (2) | 1 (0) |
| Data (2) | 0.35 (0.07) |
| Scenario (2) | 0.25 (0.21) |
| Control group (2) | 1 (0) |
| **Overall** | **0.71 (0.28)** |

For 47 of the 116 data fields extracted (41%), the 10 extractions were consistent (‘no error’). In the remaining 59% of data fields, inconsistency was either due to value errors (differences in the information returned) or voice errors (variation in how the same information was phrased or formatted), or both (see Figure 3 and supplementary Table S4 for details of error types and their acceptability).

**Figure 3.**
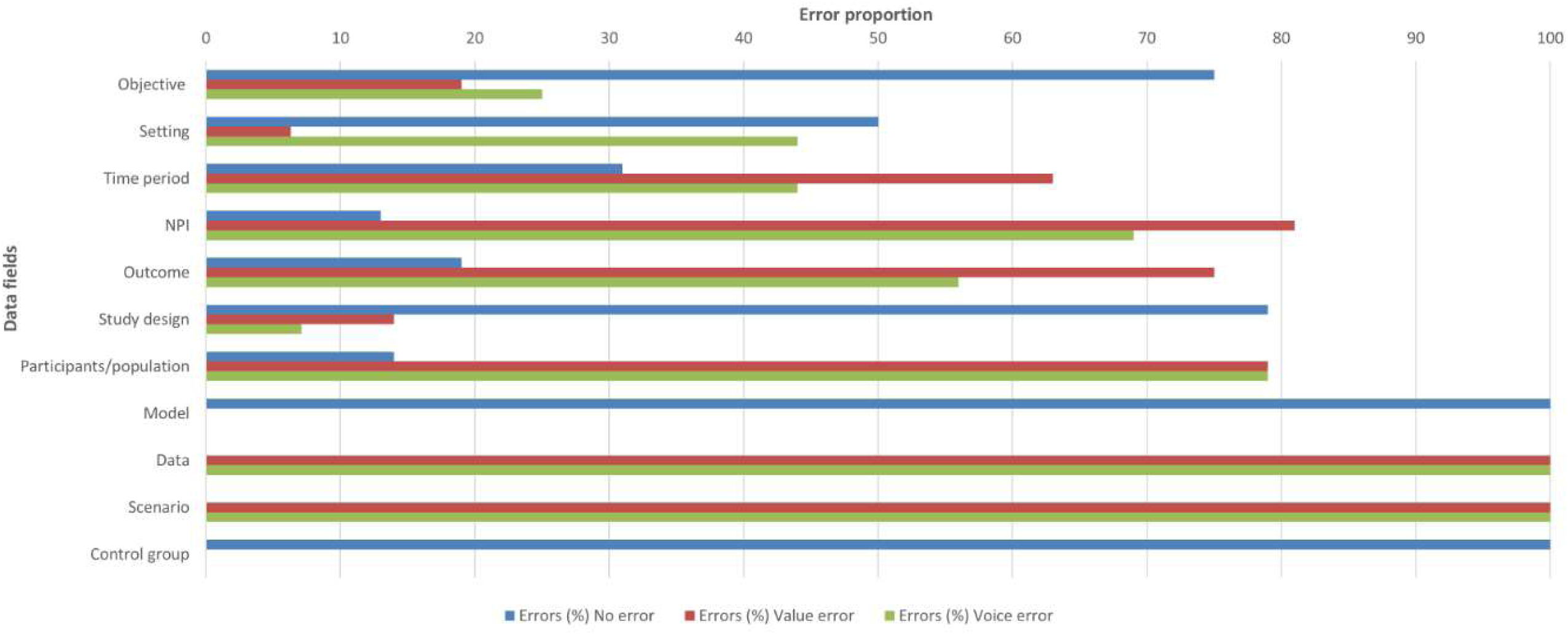
Distribution of reliability error types by data field

### 3.3 Reviewer observations of the LLM generated-extractions

Reviewers shared a number of qualitative observations related to the model-generated extractions and model behaviour. A common theme was that the model tended to be verbose, with broad and unfocused responses which gave the impression of returning all potentially relevant content from the source article, rather than summarising contextually relevant information. One reviewer described this as “responding with everything it could find, rather than answering the question”.

Recurring issues were noted across specific data fields, such as ‘time period’, ‘outcomes’ and ’NPI’s. For example, for ‘time period’, within the 46% of unacceptable extractions, the model often returned metadata such as the manuscript submission or acceptance dates, rather than the actual study period. For ‘outcomes’ and ‘NPI’s (that were rated as not acceptable for at least 50% of extractions), the model sometimes extracted content from the introduction, discussion, or title, which often referenced broader or future directions, rather than the specific interventions or outcomes assessed in the study, suggesting a potential lack of contextual prioritisation.

Reviewers also noted that prompt design appeared to influence the model’s behaviour and outputs in unintended ways. For example, the prompt for the ‘participant/population’ data field included an instruction to describe any relevant subgroups. In response, the model sometimes inferred subgroups even when none were described in the study, suggesting that the wording of prompts may have encouraged overinterpretation rather than strict extraction.

Despite these observations, reviewers noted the potential benefits of using the model as one reviewer whilst keeping a human in the loop as second reviewer. For instance, in a rapid review context, the model could act as a first reviewer and the human reviewer as a second reviewer, checking the extraction of the model. In a systematic review context, extraction by both the model and the human would be done independently and then compared to reach consensus. However, the length and complexity of outputs may limit the value. As one reviewer remarked, “it would be just as easy to read the original article”.

## 4. Discussion

This study explored the feasibility of using a bespoke RAG-LLM technical pipeline using LLaMA-3 to complete data extraction for evidence reviews in the public health context, with the intention that outputs would be quality assured by a second human reviewer (Figure 4). When extracting data from full-text articles for a range of data fields, the model returned accurate outputs (no error) for only 42% of data fields, whilst a further 28% had only minor errors. In terms of acceptability, 68% of the outputs assessed for accuracy were deemed acceptable, meaning that it would be deemed to be acceptable data extraction from a human reviewer and therefore usable in a real-world evidence review. Accuracy and reliability varied by data field, similar to previous observations by others (42). The model demonstrated strong performance for certain data fields (‘study objective’, ‘study design’ and ‘setting’), where acceptability of accuracy extractions was at least 90%, and reliability (consistency) was highest (mean maximum agreement rate of at least 0.86). However, other data fields such as ‘outcomes’, ‘participants/population’ and ‘time period’, proved more difficult for the model to extract accurately (accuracy 4.2% to 29%, accuracy acceptability 50% to 64%) and reliably (mean maximum agreement rates 0.52 to 0.69), possibly due to varied expression of data across studies and the reporting format, such as tables, or its location (43).

**Figure 4.**
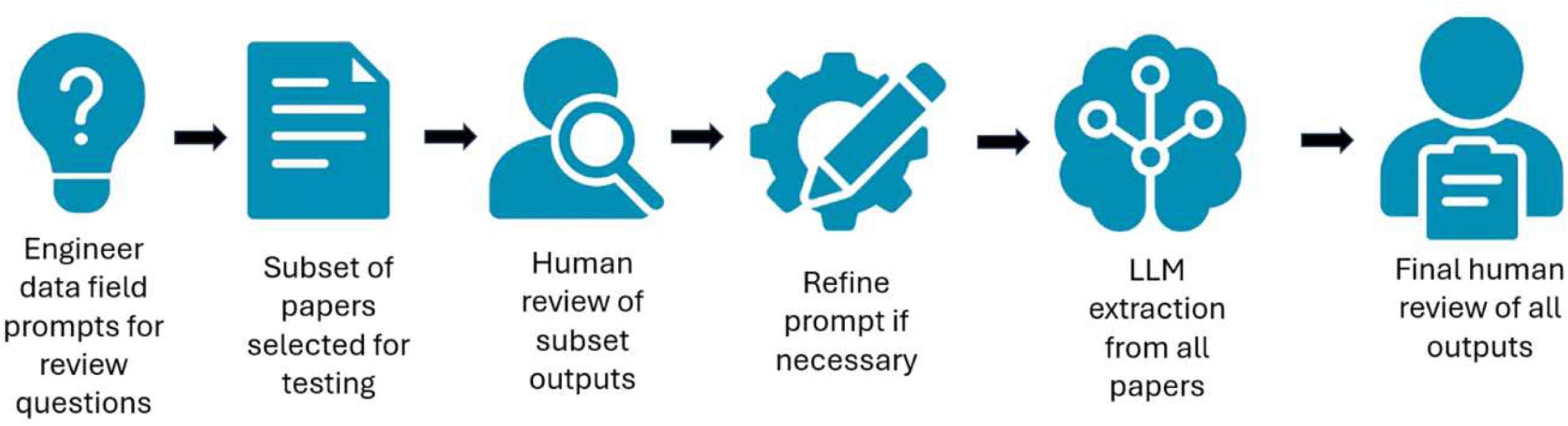
Workflow for prompt development, model extraction, and human review

Direct comparison between the findings of this evaluation and other published evaluations of LLMs’ data extraction, which report accuracy of between 68% and 96% (8–10, 14, 42), is not possible due to differences in the study designs and data fields extracted, and in the LLMs and evaluation frameworks used. In particular, the published evaluations tend to focus on extraction of data from RCTs of clinical evidence, therefore mainly numerical data, and following standardised guidance for reporting (8–10, 14). In contrast, this study evaluated the model’s performance across a range of study designs assessing public health evidence which tends to be more wordy and less structured than RCTs of clinical evidence. In this context, it is worth noting that the data extraction for the 2 RCTs included in this evaluation had the lowest acceptability of all the study designs included, which may reflect the complexity of the interventions and comparisons (both evaluated test and release strategies which involve multi-step conditional interventions) (36, 39). The range of study designs in this evaluation, along with variation in how data is reported across full texts, contributed to an increased complexity of the task but is reflective of the real-world challenges in evidence reviews in public health.

The results may reflect not only the complexity of the data fields themselves, but also the performance of the underlying model and the effectiveness of the prompt engineering approach. Performance of different LLMs varies (43, 44). Llama-3 has previously been identified as higher-performing for classification or data extraction-related tasks in the public health context (43), which informed the choice of Llama-3 for this bespoke pipeline. The effectiveness of prompt engineering also likely influenced performance (45, 46), particularly the ability of the model to extract nuanced information across data fields and from different study designs.

The reliability results raise an important question around the validity of deriving accuracy from an evaluation of a single run, which may be inadequate to evaluate model performance. This evaluation, like others (42), showed that generative AI models can produce different outputs from the same input, which complicates the interpretation of accuracy metrics. Setting the temperature close to but not at zero may have resulted in some of the variation seen, although variation has been observed with the temperature set at zero (42).

Criteria used to evaluate accuracy and reliability of LLM extractions differ in published evaluations (8–10, 14, 42). The evaluation framework used in this study was composed of a structured accuracy and reliability error classification system, and reviewer- informed acceptability judgement. Considering acceptability facilitated a more nuanced and practical assessment of model performance than binary accuracy scoring alone, by evaluating the output in the context of what a human reviewer might reasonably extract. This layered evaluation approach reflects how these tools might be used in real-world scenarios where human oversight is retained, and was aligned with the potential integration of this pipeline into rapid systematic methodologies, where a human reviewer would quality assure all model-generated extractions. However, a clearer understanding of what constitutes acceptable accuracy and reliability of LLM extractions for integration into evidence review processes is needed. Establishing what minimum threshold of acceptability is needed for the adoption of LLMs remains an open question for future research. The development of standards to assess consistency, potentially through averaged outputs, confidence thresholds, or stability metrics, will be important for the responsible adoption of AI-assisted tools in evidence review methods.

### Limitations of this work

Conclusions about differential model performance by study design cannot be drawn due to the small number of studies representing each study design within the sample. Three of the 24 studies were used during prompt development, which may have introduced an unknown degree of bias in model performance during evaluation. These studies were retained as part of the evaluation, which prioritised learning about feasibility and model behaviour over a strict separation of the data used to develop the prompts and pipeline from the data used to evaluate the model.

All data fields were weighted equally and did not account for relative differences in complexity or the potential influence of certain data fields on the interpretation of the review findings. A single set of prompts (tailored to studies included in this evaluation) and a single dataset were used, so further testing is needed to understand how performance might vary across review types, prompt phrasing, or model configurations.

As this evaluation compared LLM data extraction with extraction previously completed by humans, data was not available to compare the time impact of LLM and human data extraction.

### Next steps

With continued close collaboration between data scientists and evidence reviewers, the technical pipeline will be refined and reassessed to improve performance by exploring adjustments to the current RAG-based approach, and by testing alternative prompting methods that do not rely on retrieval components. Consideration will be given to which type of reviews, study designs, and data fields the pipeline is most appropriate and valuable for. Time savings and resource impact will also need to be captured in future testing.

It will be important to consider the principles and practicalities of responsible AI integration and develop appropriate guidelines for its use. This includes ensuring transparency in how technical pipelines and the models embedded within them are developed, evaluated, and deployed. Users will need to maintain transparent documentation of prompt engineering, model parameters, and versioning, and establish clear decision thresholds and audit trails to support reproducibility and accountability.

There also needs to be a consensus-based decision about acceptable performance thresholds and risk. These considerations are aligned with guidance on responsible AI use in evidence synthesis, currently in development (47), which highlights the importance of fairness, robustness, and transparency in the application of AI in evidence reviews.

## 5. Conclusion

This evaluation demonstrates the potential for LLMs, when paired with human quality assurance, to support data extraction in evidence reviews that include a range of study designs, however further improvements in performance are required before the model can be introduced into review workflows. While overall accuracy was modest, acceptability of outputs, defined as their practical usability by reviewers was higher, showing potential for real-world application. Performance varied across data fields, and reliability issues highlighted the limitations of single run evaluations. However, despite overall variability, the model performed well for the data fields ‘objective’, ‘setting’, and ‘study design’. The layered evaluation framework, which considered acceptability as well as accuracy and reliability, offered a more nuanced measure of how these tools might function in evidence review contexts, and the co-development process between data scientists and evidence reviewers was important in aligning technical design with practical needs. However, further testing across review types, refined prompting strategies, and clearer thresholds for acceptable model performance are needed.

Future work must also address the principles of responsible AI integration, including transparency, reproducibility, and appropriate risk-benefit trade-offs. With these considerations in place, semi-automated data extraction using LLMs could play a valuable role in improving the efficiency of public health evidence reviews.

## Data Availability

All data produced are available in supplementary files 2 and 3

## Acknowledgements

We acknowledge the support of colleagues in UKHSA Chief Data Officer Group (Chris Moyle, Josh Harris, Luke Hounsome, Timothy Laurence, Leo Loman), in UKHSA Knowledge and Library Services, and in UKHSA Evidence Network.

## Disclaimer

The views expressed in this article are those of the authors and are not necessarily those of UK Health Security Agency or the Department of Health and Social Care.

