## Supplementary file 1 for "Assessing the feasibility and acceptability of a bespoke large language model pipeline to extract data from different study designs for public health evidence reviews"

**Table S1. Example LLM output**

| **Study design** | **Outcome** | **NPI** | **Setting** | **Participants** | **Study period** | **Objective** |
|---|---|---|---|---|---|---|
| The study design was: A mixed-methods study, including a survey, interviews, and focus groups. | The main outcomes measured in the study were:<br>- PCR testing completion<br>- Gender as a dummy coded variable<br>- Covariates (gender and year of study) | There was no non-pharmaceutical interventions mentioned in the chunks provided. | The setting of the study was at the University of Nottingham, where they conducted a mixed-methods study on the perceptions and experiences of the SARS-CoV-2 asymptomatic testing service. | The study involved:<br>- 41 individuals who attended interviews or focus groups, including 31 students (21 first year, 10 final year) | The duration of the study was:<br>- 16 days | The main objective of the study was:<br>- To investigate the perceptions and experiences of the University of Nottingham Pilot SARS-CoV-2 Asymptomatic Testing Service |

**Table S2. System message and prompt used for LLama 3-70b**

**System message:** "You are a helpful, respectful and honest assistant. Always answer as helpfully as possible, while being safe. Your role is to take sentences from academic papers and summarise their information. Extract and summarise the information using only the given question, chunks, codes and the study title delimited by ###. Do not return any information found in the examples below which is delimited by <example> </example>. Use the study title as a guide when extracting and summarising the information from the chunks. "

**Prompt:** """

Your task is to perform data extraction on studies pertaining to COVID-19 vaccination and transmission which will be provided to you in chunks. Your goal is to extract relevant data from these studies.

Your data extraction should be limited to the content available in the chunks, and no external sources should be referred to.

Output the information in bullet points, and as concisely as possible.

Only return dates if the question contains a measure of time.

Let's think through this carefully, step by step. Then, identify which response is the best answer to the question asked.

Here is an example :

<example>

Topic: study design

Question: What was the study design?

Study Title: Randomised Controlled Trial of a New Drug for Hypertension

Chunks: ```This study was a double blinded, placebo-controlled randomised controlled trial based on 200 participants in England ```

Response: The study design was:

- Double-blind, placebo-controlled randomised controlled trial

</example>

###

Topic: {topic}

Question: {additional_info}

Study Title: {title}

"""

**Table S3. Acceptability of accuracy extractions by study design**

| Data field | **RCTs** (2 studies, 16 data fields) | **Prospective longitudinal** (5 studies, 35 data fields) | **Cross-sectional** (3 studies, 21 data fields | **Ecological** (4 studies, 28 data fields) | **Mixed-methods** (4 studies, 28 data fields) | **Qualitative** (3 studies, 21 data fields) | **Modelling** (3 studies, 24 data fields) | **All study designs (24 studies, 173 data fields)** |
|---|---|---|---|---|---|---|---|---|
| Objective | 2 (100%) | 5 (100%) | 3 (100%) | 4 (100%) | 3 (75%) | 2 (67%) | 3 (100%) | 22 (92%) |
| Setting | 2 (100%) | 5 (100%) | 3 (100%) | 4 (100%) | 3 (75%) | 2 (67%) | 3 (100%) | 22 (92%) |
| Time period | 1 (50%) | 2 (40%) | 3 (100%) | 3 (75%) | 1 (25%) | 1 (33%) | 2 (67%) | 13 (54%) |
| NPI | 0 (0%) | 1 (20%) | 3 (100%) | 1 (25%) | 3 (75%) | 1 (33%) | 2 (67%) | 11 (46%) |
| Outcome | 1 (50%) | 2 (40%) | 2 (67%) | 2 (50%) | 1 (25%) | 2 (67%) | 2 (67%) | 12 (50%) |
| Study design | 2 (100%) | 4 (80%) | 2 (67%) | 4 (100%) | 4 (100%) | 3 (100%) | NA | 19 (90%) |
| Participants/ population | 0 (0%) | 4 (80%) | 2 (67%) | 4 (100%) | 2 (50%) | 2 (67%) | NA | 14 (67%) |
| Model | NA | NA | NA | NA | NA | NA | 3 (100%) | 3 (100%) |
| Data | NA | NA | NA | NA | NA | NA | 1 (33%) | 1 (33%) |
| Scenario | NA | NA | NA | NA | NA | NA | 0 (0%) | 0 (0%) |
| Control group | 1 (50%) | NA | NA | NA | NA | NA | NA | 1 (50%) |
| **Totals** | **9 (56%)** | **23 (66%)** | **18 (86%)** | **22 (79%)** | **17 (61%)** | **13 (62%)** | **16 (67%)** | **118 (68%)** |

NA = not applicable (data field not extracted for the study design)

**Table S4 Reliability errors and acceptability by data field (total number of errors = 157)**

| **Data field (number of studies)** | **Value error (%)** | **Voice error (%)** | **No error (%)** | **Value not returned (%)** | **Acceptability of errors (% acceptable)** |
|---|---|---|---|---|---|
| Objective (16) | 3 (19%) | 4 (25%) | 12 (75%) | 0 | 84% |
| Setting (16) | 1 (6.3%) | 7 (44%) | 8 (50%) | 0 | 94% |
| Time period (16) | 10 (63%) | 7 (44%) | 5 (31%) | 0 | 68% |
| NPI (16) | 13 (81%) | 11 (69%) | 2 (13%) | 0 | 54% |
| Outcome (16) | 12 (75%) | 9 (56%) | 3 (19%) | 0 | 58% |
| Study design (14) | 2 (14%) | 1 (7.1%) | 11 (79%) | 0 | 93% |
| Participants/ population (14) | 11 (79%) | 11 (79%) | 2 (14%) | 0 | 58% |
| Model (2) | 0 | 0 | 2 (100%) | 0 | 100% |
| Data (2) | 2 (100%) | 2 (100%) | 0 | 0 | 50% |
| Scenario (2) | 2 (100%) | 2 (100%) | 0 | 0 | 50% |
| Control group (2) | 0 | 0 | 2 (100%) | 0 | 100% |
| **Totals** | **56*** | **54*** | **47*** | **0** | **69%** |

*47 of 116 data fields (41%) had no errors. The remaining 69 (59%) data fields had a voice and/or value error (total 56 value errors, 54 voice errors)
